# An outpatient integrated management program reduced the need for hospitalizations of CKD patients: findings from the PIRP project

**DOI:** 10.1101/2025.02.19.25322544

**Authors:** Antonio Santoro, Dino Gibertoni, Vittorio Albertazzi, Andrea Buscaroli, Simonetta Cimino, Gabriele Donati, Enrico Fiaccadori, Mariacristina Gregorini, Gaetano La Manna, Emanuele Mambelli, Renato Rapanà, Roberto Scarpioni, Alda Storari, Annalisa Zucchelli, Marcora Mandreoli

## Abstract

**Background and hypothesis:** In patients with moderate or severe renal disease, hospitalization is often required because of poorly controlled co-morbidities. We aimed to provide evidence that an outpatient health program involving a close collaboration between nephrologists and General Practitioners can be successful in reducing hospitalizations in non-dialysis chronic kidney disease patients.

**Methods:** Observational cohort study on 17,036 stage 1-5 chronic kidney disease patients enrolled in the Emilia-Romagna (Italy) PIRP project between 1st April 2004 and 31st December 2015, and their 70,560 hospitalizations registered in the four years preceding and following their enrolment in the project. Interrupted Time Series analysis was used to estimate hospitalizations’ trend summarized on 4-monthly basis.

**Results:** Among patients who survived 4 years in non-dialysis chronic kidney disease condition, a 2.9% reduction in hospitalizations was observed in the four years following the enrolment in PIRP compared to the four years previously. The change in hospitalizations’ trend was estimated at −8.09 admission per 1,000 patients and 4-month period. This decrease was mainly accountable to hospitalizations whose main diagnoses at discharge were diseases of the circulatory system and the genitourinary system (−2.68 and −4.76 admissions per 1,000 patients respectively). Patients with heart failure and those with coronary artery disease displayed large reductions in hospitalization trend (−17.08 and −9.48 admissions per 1,000 patients respectively). A reduction of hospitalizations with similar magnitude was also observed for the advanced stages of CKD.

**Conclusion:** The implementation of an integrated public health project that provides for the early management and continuity of care of CKD patients may be a way to reduce hospitalizations, particularly those related to cardiovascular and genitourinary diagnoses.

**Key learning points:** *What was known:* With an integrated and structured program based on the collaboration between nephrologists and General Practitioners, it is possible to better control progression and comorbidity in CKD.

*This study adds:* By continuously following CKD patients with a close collaboration between nephrologists and General Practitioners, hospitalizations can be reduced.

*Potential impact:* The implementation of an integrated model of outpatient management of CKD patients like the PIRP might be beneficial also on hospitals’ organization and costs, and ultimately on patients’ quality of life.

**Graphical abstract:** 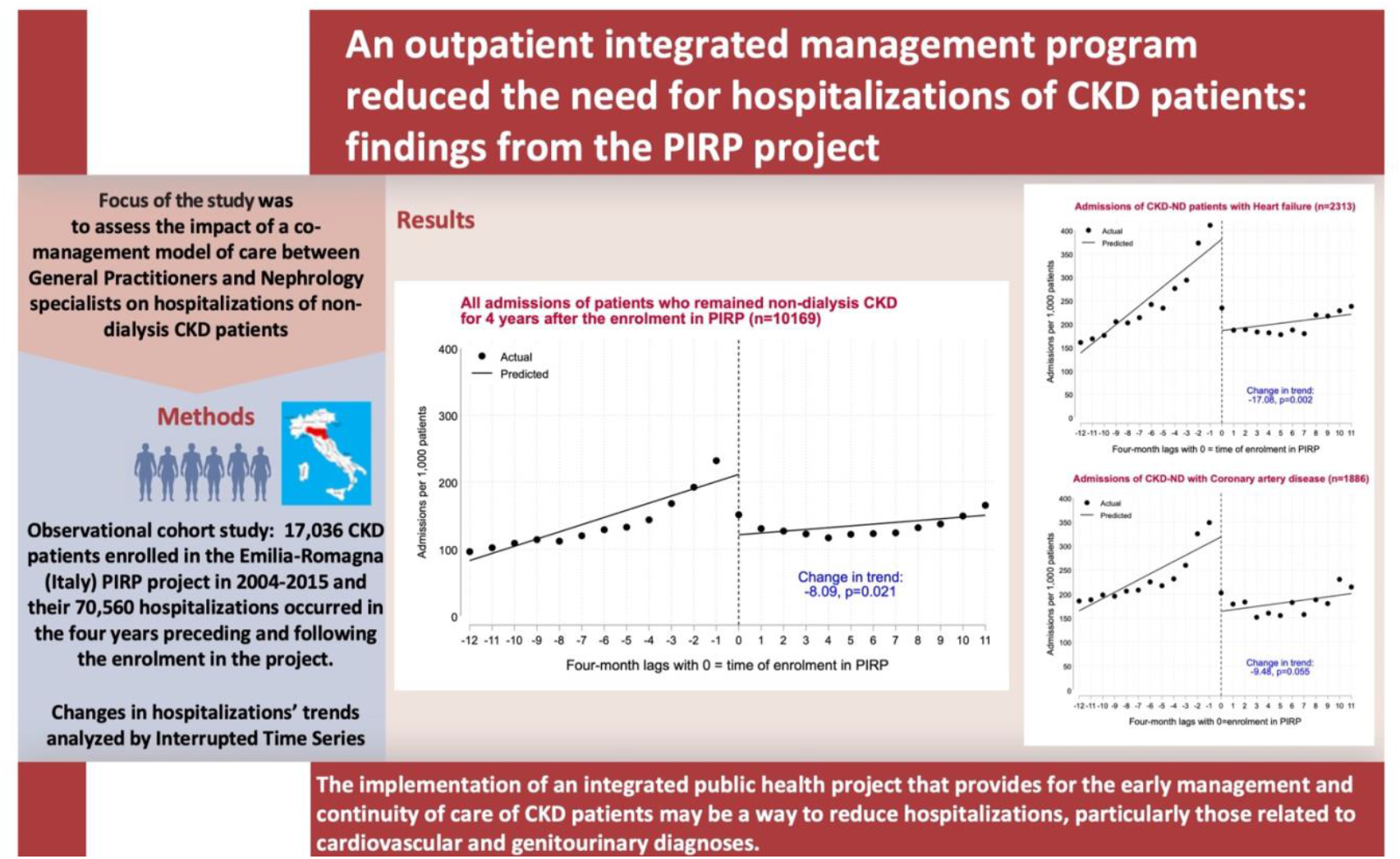

## INTRODUCTION

Hospitalization is frequent during the course of chronic kidney disease (CKD) under the burden of co-morbidities and peaks in the months immediately before and after the initiation of dialysis [1]. The most frequent causes of admission are those for cardiovascular diseases [2–5], infections [6] and kidney events [2–3]. A recent study by Chong et al. reports that in CKD patients acute care utilization was 3-8 times higher than in the general population [7]. Diwan et al. found that non-dialysis CKD (CKD-ND) patients had 1.7 admissions per person-year, 8 times higher than the general population [8]. Early and adequate interventions are essential to delay or prevent the progression of kidney disease [9]. A successful intervention in reducing CKD progression should also entail a reduction in the number of hospitalizations, but this needs to be demonstrated. In the Emilia-Romagna north-eastern region of Italy, the PIRP (Prevenzione dell’Insufficienza Renale Progressiva) project was established in 2004, with the aims of timely detecting patients with CKD, and to prevent or delay end-stage kidney disease by the implementation of a dedicated care pathway in which General Practitioners (GPs) and hospital nephrologists from all the 13 regional hospital units cooperate. The outcomes achieved by PIRP have already been described, in terms of reducing estimated glomerular filtration rate (eGFR) decline and the number per year of patients who are forced to start dialysis [10–11].

The aim of the present study was to verify whether the PIRP project could have an additional positive impact by reducing the number of hospitalizations in CKD-ND patients. To address this, a comparison between the hospitalization trends in the 4 years preceding each patient’s enrolment in PIRP and 4 years after PIRP enrolment was made.

## MATERIALS AND METHODS

### The PIRP Project

The PIRP project started in 2004 to implement a unified regional care pathway targeting the growing number of CKD patients. It is a clinical project that provides for a close collaboration between GPs and nephrologists in order to obtain an early referral to specialists [10–11].

After the initial phase of training of GPs, specifically aimed towards the early recognition of people with kidney disease and adoption of the most appropriate therapies, the project envisaged the opening of nephrological outpatient clinics specifically dedicated to patients with CKD. Patients entering this project have an easy access to the care path and are treated jointly by GPs and nephrologists with pharmacological and nonpharmacological strategies according to recommendations from CKD Guidelines. The process of care includes medication reconciliation to support the appropriate drug dosing and avoidance of nephrotoxicity. Another important task of the project is to identify patients at high risk of progression towards advanced renal failure and to introduce them to a more stringent follow-up program [12]. Details regarding patients’ management are shown in Table 1. After each outpatient visit, the clinical and laboratory parameters of each patient are routinely entered into a web-based database (PIRP Registry). The project is still active and as of June 30th, 2024, its registry includes data from 163,173 visits of 36,373 patients with median follow-up of 45 months.

**Table 1:**
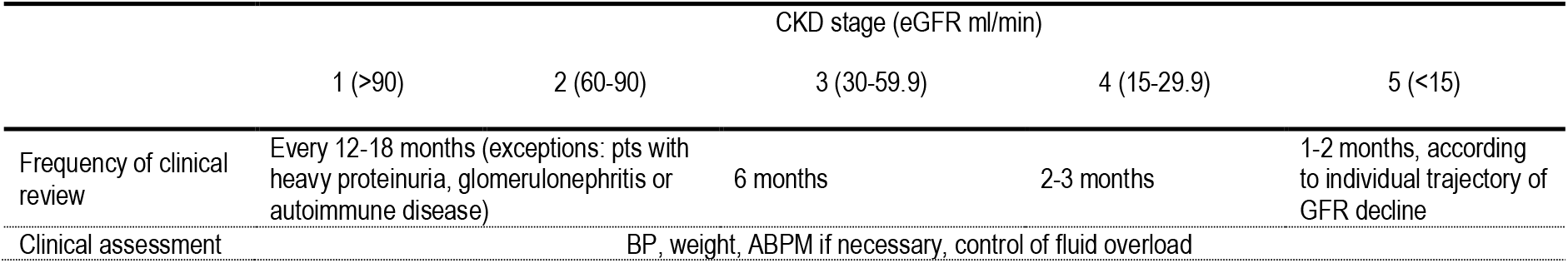
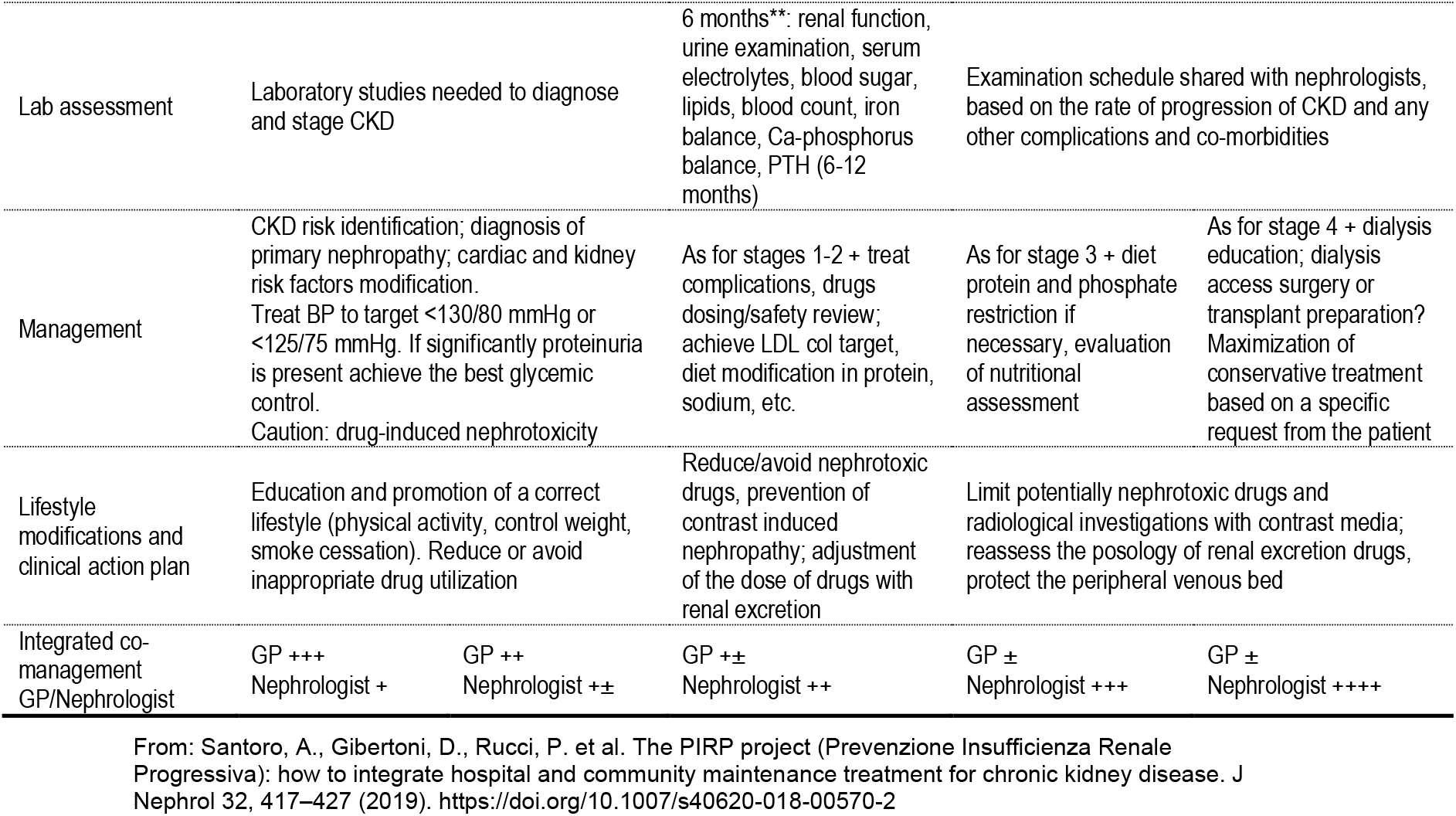
CKD Management according to stage.

### Study population

Eligible patients for the present study were those enrolled in the PIRP project between 1.4.2004 and 31.12.2015 and resident in the Emilia-Romagna region (98.5% of all PIRP patients). Exclusion criteria were being lost to follow-up before 4 years since enrolment, and lacking correct identifier data to link with regional registries (Fig.1).

**Figure 1:**
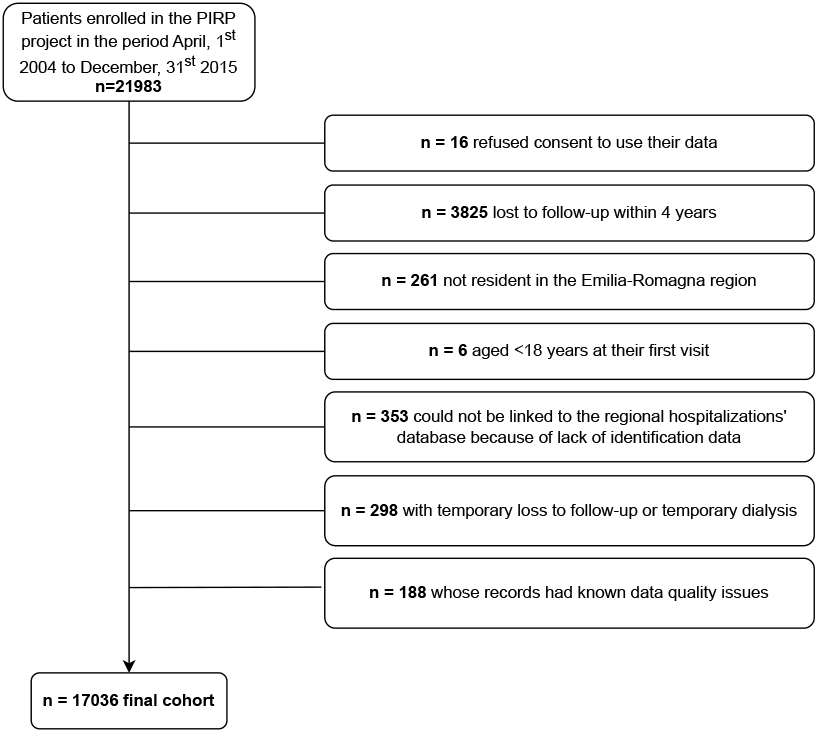
Flow chart describing the derivation of the study cohort.

Ethical approval to conduct this retrospective research was granted by the Area Vasta Emilia Centro (AVEC) Ethics Committee, approval identification No. 341/2022/Oss/AOUBo.

### Data sources

Alongside with demographic, anamnestic and laboratory data, the PIRP registry periodically receives data from the regional registries of hospital admissions and mortality, by means of a deterministic linkage through the pseudonymized regional identifier of patients. Data used for the present study included all hospitalizations recorded from 1^st^ April 2000 to 31^st^ December 2019, from which admission and discharge dates and the related wards, main diagnoses at admission (coded using ICD9-CM classification and grouped with criteria given in the Supplementary Materials) and discharge modality were retrieved. The diagnoses of hospitalizations prior to patients’ entry in PIRP were also used to calculate the Charlson Comorbidity Index (CCI) and to identify patients with heart failure (HF) and with coronary artery disease. The mortality registry feeds the PIRP database with date and cause of death. A deterministic linkage with the regional database of dispensed drugs was specifically created for this study.

### Statistical analysis

Laboratory data were those recorded at the first PIRP visit or within one year from the first visit if the baseline data was missing. eGFR was estimated using data from the most recent laboratory examination with respect to the first PIRP visit by the 2009 CKD Epidemiology Collaboration (CKD-EPI) creatinine equation [13]. Since 2009, calibrated and IDMS traceable serum creatinine method [14] is in use.

Patient characteristics were summarized using absolute frequencies, percentages, or mean+standard deviation and compared among patients who survived, those who died and those who reached ESKD during the first 4 years after enrolment in PIRP. Continuous variables were compared using ANOVA, while categorical variables were compared using the likelihood-ratio χ2 test.

The Interrupted Time Series (ITS) was used to evaluate time series of hospitalization over a time frame that includes the inception of an intervention which is presumed to modify the observed trend. With ITS, three parameters of the time series under study are estimated: the linear slope observed before the intervention, the change in level occurred at the first observation after the intervention, and the change in slope between the periods before and after the intervention. The hypotheses of invariance in level and in trend after the intervention are based on the counterfactual that in the absence of intervention the trend would have continued the same [15]. Prais-Winsten estimators were used, assuming that errors would follow a first-order autoregressive process. In our study, the intervention was CKD-ND patients’ enrolment in the PIRP project, and the outcome was the number of hospital admissions per 1,000 patients, summarized in each four-month period in the four years preceding and following the first visit in PIRP. The four-year length observation period following enrolment in PIRP allowed to carry out ITS with the minimum recommended number of timepoints [16] and was deemed adequate to display the potential impact of the intervention on hospitalizations. A period of the same length preceding the intervention was chosen to balance the time series with respect to the intervention. Hospitalizations assignment to a time period was made according to the admission date.

As patients’ selection span on a 12-year long period during which policies on hospitalizations might have changed, sensitivity analyses were conducted on 3-year subsets, to verify the consistency of findings over time. The analyses were conducted on the whole study population and on subgroups defined by patients’ clinical outcome, main diagnosis at hospital admission, CKD stage, history of heart failure (HF) and coronary artery disease (CAD).

The proportions of patients who were already under treatment for the main ATC groups of interest for CKD before entering the PIRP project were also calculated, along with the number of patients whose treatment was modified during the first year in PIRP, either by introducing drugs or discarding them.

All analyses were carried out using Stata v.18.0 and with statistical significance set at p<0.05. Specifically, the user-written itsa procedure [17] was used for ITS analysis.

## RESULTS

Patients eligible for participating in the study were n=21,983. At the end of the selection process a final cohort of 17,036 patients with data on 70,560 hospitalizations with admissions dating from 2^nd^ May 2000 to 19^th^ November 2019 was established. Patients who reached 4 years follow-up in the PIRP project (hereafter referred as “stayers”) were 10,169 (59.7%), while 5,047 (29.6%) died and 1,820 (10.7%) started renal replacement therapy (hereafter referred as ESKD) within 4 years (Table 2). Almost all clinical characteristics differed significantly among stayers, died and ESKD patients. Notably, stayers had higher levels of eGFR and hemoglobin, and a lower Charlson Comorbidity Index; patients who died were older and had higher prevalence of HF and CAD; those who reached ESKD were younger and had a higher prevalence of proteinuria.

**Table 2:** Characteristics of the study population at the first visit in the PIRP project by status after 4 years.

|  | All<br>n=17,036 | Stayers<br>n=10,169 | Non-stayers<br>n=6,867 | Died<br>n=5,047 | ESKD<br>n=1,820 | Test (*) |
| --- | --- | --- | --- | --- | --- | --- |
| Age (years) | 73.1±11.9 | 71.0±11.8 | 76.2±11.4 | 79.5±8.2 | 67.1±13.9 | <0.001 |
| Females | 5,941 (34.9%) | 3,493 (34.3%) | 2,448 (35.6%) | 1,805 (35.8%) | 643 (35.3%) | 0.218 |
| Smoking status |  |  |  |  |  | <0.001 |
| Ex-smoker | 4,660 (40.6%) | 2,801 (40.6%) | 1,859 (40.6%) | 1,470 (43.3%) | 389 (33.0%) |  |
| Smoker | 1,154 (10.1%) | 711 (10.3%) | 443 (9.7%) | 248 (7.3%) | 195 (16.5%) |  |
| Non-smoker | 5,655 (49.3%) | 3,383 (49.1%) | 2,272 (49.7%) | 1,676 (49.4%) | 596 (50.5%) |  |
| Patient with proteinuria (yes/no) | 5,982 (54.4%) | 2,655 (39.2%) | 2,356 (55.8%) | 1,307 (44.1%) | 1,049 (83.5%) | <0.001 |
| Type 2 diabetes (yes/no) | 6,931 (40.7%) | 3,994 (39.3%) | 2,937 (42.8%) | 2,148 (42.6%) | 789 (43.4%) | <0.001 |
| Main CKD diagnosis |  |  |  |  |  | <0.001 |
| Diabetic nephropathy | 2,200 (12.9%) | 1,138 (11.2%) | 1,062 (15.5%) | 684 (13.6%) | 378 (20.8%) |  |
| Glomerulonephritis | 785 (4.6%) | 522 (5.1%) | 263 (3.8%) | 80 (1.6%) | 183 (10.1%) |  |
| Hypertensive nephropathy | 9,889 (58.0%) | 5,945 (58.5%) | 3,944 (57.4%) | 3,195 (63.3%) | 749 (41.2%) |  |
| Pyelonephritis, interstitial nephrop. | 909 (5.3%) | 584 (5.7%) | 325 (4.7%) | 222 (4.4%) | 103 (5.7%) |  |
| Polycystic kidney | 472 (2.8%) | 293 (2.9%) | 179 (2.6%) | 44 (0.9%) | 135 (7.4%) |  |
| Single kidney | 642 (3.8%) | 462 (4.5%) | 180 (2.6%) | 144 (2.9%) | 36 (2.0%) |  |
| Unknown cause | 1,892 (11.1%) | 1,064 (10.5%) | 828 (12.1%) | 629 (12.5%) | 199 (10.9%) |  |
| Other nephropathies | 247 (1.4%) | 161 (1.6%) | 86 (1.3%) | 49 (1.0%) | 37 (2.0%) |  |
| Charlson Comorbidity Index | 3.43±2.87 | 2.59±2.48 | 4.68±2.95 | 4.96±3.05 | 3.89±2.51 | <0.001 |
| Heart failure (yes/no) | 5,408 (31.7%) | 2,313 (22.7%) | 3,095 (45.1%) | 2,542 (50.4%) | 553 (30.4%) | <0.001 |
| Coronary artery disease (yes/no) | 3,694 (21.7%) | 1,886 (18.5%) | 1,808 (26.3%) | 1,480 (29.3%) | 328 (18.0%) | <0.001 |
| eGFR (ml/min/1.73 m<sup>2</sup>) | 32.11 ±14.54 | 36.55 ±14.57 | 25.51 ±11.70 | 27.78 ±11.45 | 19.19 ±9.96 | <0.001 |
| BMI (kg/m<sup>2</sup>) | 27.27 ±4.82 | 27.64 ±4.73 | 26.72 ±4.91 | 26.57 ±4.83 | 27.12 ±5.11 | <0.001 |
| Hemoglobin | 12.34 ±1.84 | 12.79 ±1.78 | 11.67 ±1.72 | 11.67 ±1.76 | 11.66 ±1.62 | <0.001 |
| Creatinine | 2.17 ±0.95 | 1.88 ±0.64 | 2.59 ±1.16 | 2.27 ±0.85 | 3.48 ±1.42 | <0.001 |
| Total cholesterol | 185.95 ±44.71 | 189.30 ±43.15 | 180.41 ±46.65 | 177.65 ±44.14 | 187.29 ±51.78 | <0.001 |
| HDL cholesterol | 49.03 ±15.17 | 49.75 ±15.24 | 47.79 ±14.96 | 47.67 ±15.07 | 48.10 ±14.68 | <0.001 |
| LDL cholesterol | 108.27 ±38.99 | 110.33 ±38.63 | 104.72 ±39.34 | 102.89 ±37.08 | 109.61 ±44.50 | <0.001 |
| Triglycerides | 149.24 ±88.03 | 151.79 ±89.95 | 145.05 ±84.62 | 139.49 ±78.72 | 158.66 ±96.26 | <0.001 |
| Uric acid | 6.49 ±1.87 | 6.48 ±1.75 | 6.51 ±2.05 | 6.61 ±2.15 | 6.27 ±1.73 | <0.001 |
| Potassium | 4.71 ±0.57 | 4.68 ±0.54 | 4.75 ±0.61 | 4.72 ±0.59 | 4.84 ±0.63 | <0.001 |
| Phosphates | 3.67 ±0.81 | 3.49 ±0.68 | 3.92 ±0.89 | 3.71 ±0.77 | 4.33 ±0.98 | <0.001 |
| Glycated hemoglobin (mmol) | 53.25 ±17.82 | 53.32 ±17.61 | 53.14 ±18.15 | 53.66 ±18.11 | 51.81 ±18.18 | 0.165 |

(\*) test of the hypothesis of equality among survivors, died and ESKD (ANOVA for continuous variables, chi-square for categorical variables)

Among stayers, patients never hospitalized increased by 10.9% (from 3,723 to 4,130) in the 4 years following enrolment in the PIRP project compared to the previous 4 years and included 2,248 patients (13.2%) who were never hospitalized in the 8 years of observation. The overall number of hospital admissions of stayers decreased by 2.9% (n=495) in the 4 years after enrolment in the PIRP project (Table 3). The most frequent main hospitalization diagnosis included diseases of the circulatory system, which displayed a 15.7% reduction in admissions. Hospitalizations for diseases of the genitourinary system accounted for approximately 18% of all admissions and decreased by 4.1%, while admissions for neoplasms decreased by 21.2%. Admissions for diseases of the respiratory system, injuries and for infection diseases were less frequent but increased remarkably (+73.3%; +33.8% and +46.4% respectively).

**Table 3:** Number of hospital admissions before and after patients’ enrolment in the PIRP project, by main diagnosis at discharge –stayers (patients who survived 4 years without reaching ESKD)

| Main diagnosis groups | N. admissions<br>4 years before<br>PIRP | N. admissions<br>4 years after<br>PIRP | Diff. (%) |
| --- | --- | --- | --- |
| Diseases of the Circulatory System | 5904 | 4976 | -15.7 |
| Diseases of the Genitourinary System | 3041 | 2915 | -4.1 |
| Diseases of the Respiratory System | 846 | 1466 | +73.3 |
| Injury and Poisoning | 763 | 1021 | +33.8 |
| Diseases of the Digestive System | 1341 | 1362 | +1.6 |
| Neoplasms | 1554 | 1224 | -21.2 |
| Infectious and Parasitic Diseases | 267 | 391 | +46.4 |
| Other diagnoses | 3070 | 2936 | -4.4 |
| *Total* | *16786* | *16291* | *-2.9* |

ITS performed on hospital admissions per 1,000 patients and 4-month periods (AP4M) showed that after the intervention (i.e. the enrolment in the PIRP project) a sharp decrease in admissions’ level and trend was observed in stayers, while patients who died or started ESKD displayed an initial reduction but subsequently maintained a trend similar to the one observed before entering PIRP (Figure 2, Table 4).

**Table 4:** Results of the Interrupted Time Series analysis. Hospitalizations by 1000 patients and four-month period.

|  | Trend before PIRP |  | Change with PIRP enrolment |  | Change in trend |  | Trend during PIRP |  |
| --- | --- | --- | --- | --- | --- | --- | --- | --- |
|  | coeff. | p-value | coeff. | p-value | coeff. | p-value | coeff. | p-value |
| Stayers (n=10,169) | 10.73 | <0.001 | -90.15 | <0.001 | -8.09 | 0.021 | 2.64 | 0.191 |
| Non-stayers (n=6,867) | 22.04 | <0.001 | -76.21 | 0.025 | -1.26 | 0.869 | 20.77 | <0.001 |
| Died (n=5,047) | 21.67 | <0.001 | -55.03 | 0.119 | -3.57 | 0.581 | 18.10 | <0.001 |
| ESKD (n=1,820) | 20.52 | 0.013 | -101.23 | 0.132 | 7.05 | 0.552 | 27.58 | 0.002 |
| Stayers (n=10,169) | 10.73 | <0.001 | -90.15 | <0.001 | -8.09 | 0.021 | 2.64 | 0.191 |
| Non-stayers (n=6,867) | 22.04 | <0.001 | -76.21 | 0.025 | -1.26 | 0.869 | 20.77 | <0.001 |
| Died (n=5,047) | 21.67 | <0.001 | -55.03 | 0.119 | -3.57 | 0.581 | 18.10 | <0.001 |
| ESKD (n=1,820) | 20.52 | 0.013 | -101.23 | 0.132 | 7.05 | 0.552 | 27.58 | 0.002 |
| *By main diagnosis at discharge (stayers)* |  |  |  |  |  |  |  |  |
| Genitourinary System | 4.62 | 0.001 | -35.09 | <0.001 | -4.76 | 0.020 | -0.14 | 0.903 |
| Circulatory System | 3.49 | <0.001 | -34.79 | <0.001 | -2.68 | 0.004 | 0.81 | 0.142 |
| Neoplasms | 0.48 | 0.005 | -6.43 | <0.001 | -0.30 | 0.226 | 0.19 | 0.236 |
| Respiratory System | 0.51 | <0.001 | -1.98 | 0.084 | 0.16 | 0.308 | 0.67 | <0.001 |
| Digestive System | 0.28 | 0.002 | -3.32 | <0.001 | 0.02 | 0.868 | 0.30 | 0.001 |
| Infective diseases | 0.29 | <0.001 | -1.92 | 0.001 | -0.09 | 0.295 | 0.20 | 0.002 |
| Injury and Poisoning | 0.25 | 0.012 | -1.37 | 0.139 | 0.09 | 0.504 | 0.34 | 0.001 |
| Other diagnoses | 0.99 | 0.001 | -7.28 | 0.004 | -0.92 | 0.021 | 0.04 | 0.876 |
| *Patients with heart failure (HF)* |  |  |  |  |  |  |  |  |
| All admissions | 20.25 | <0.001 | -195.16 | <0.001 | -17.08 | 0.002 | 3.17 | 0.309 |
| Admissions with HF as main diagnosis | 4.86 | <0.001 | -45.75 | <0.001 | -3.61 | 0.004 | 1.26 | 0.098 |
| *Patients with coronary artery disease (CAD)* |  |  |  |  |  |  |  |  |
| All admissions | 12.83 | <0.001 | -154.97 | <0.001 | -9.48 | 0.055 | 3.35 | 0.267 |
| Admissions with CAD as main diagnosis | 2.35 | 0.007 | -59.69 | <0.001 | -2.68 | 0.037 | -0.32 | 0.681 |
| *By CKD stage at enrolment in PIRP* |  |  |  |  |  |  |  |  |
| CKD 1-2 | 8.81 | 0.001 | -65.95 | 0.005 | -8.24 | 0.027 | 0.57 | 0.808 |
| CKD 3A | 5.56 | <0.001 | -44.40 | <0.001 | -4.74 | <0.001 | 0.82 | 0.277 |
| CKD 3B | 7.81 | <0.001 | -65.46 | <0.001 | -5.04 | 0.021 | 2.77 | 0.046 |
| CKD 4 | 14.21 | <0.001 | -118.49 | <0.001 | -10.88 | 0.032 | 3.33 | 0.267 |
| CKD 5 | 41.27 | 0.002 | -367.02 | <0.001 | -37.06 | 0.066 | 4.21 | 0.722 |

**Figure 2:**
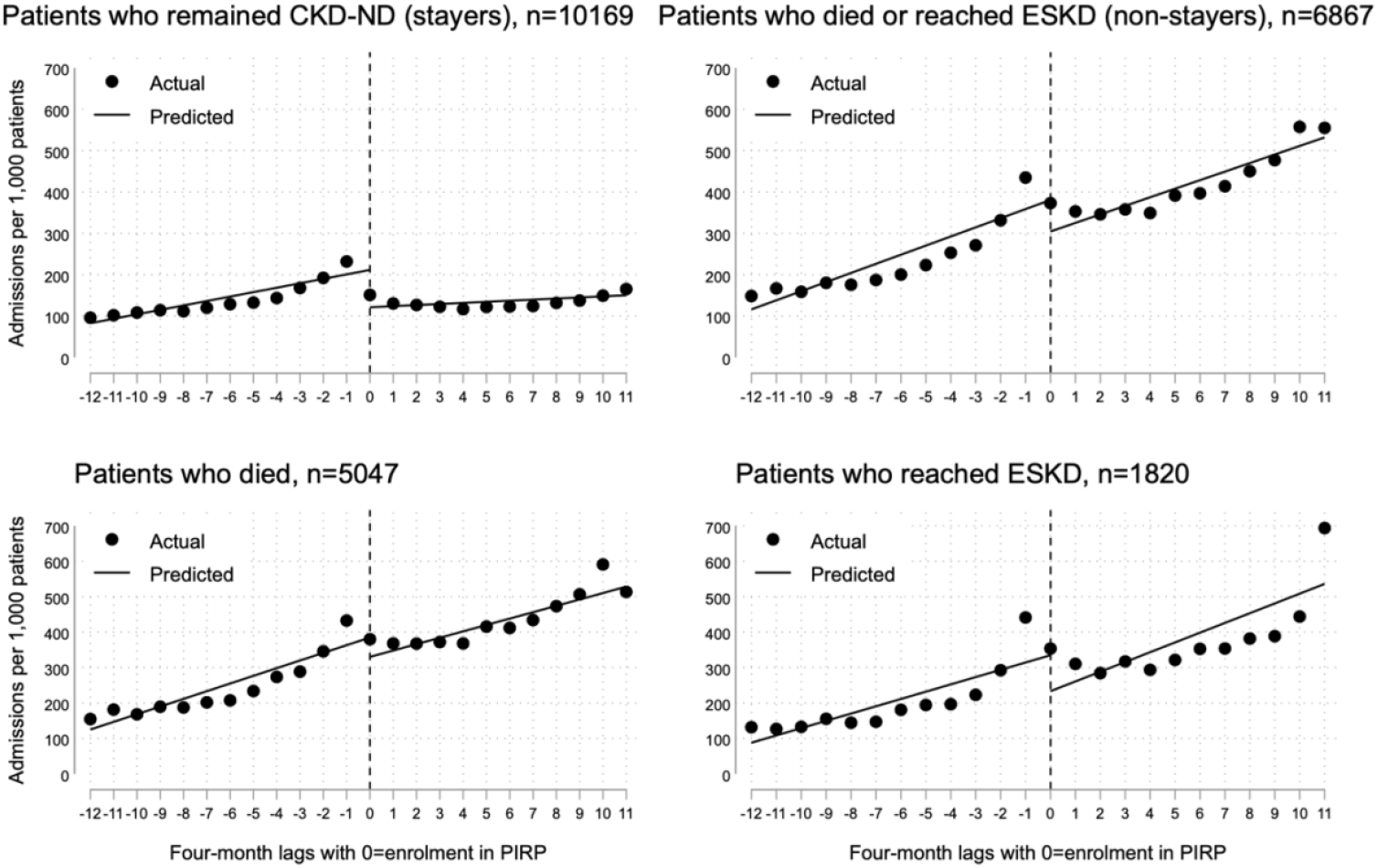
ITS of hospitalizations per 1,000 patients and four-month periods of stayer and non-stayer CKD-ND patients. Note: each point represents the number of admissions per 1,000 patients per four-months; on the X-axis, timepoint 0 corresponds to the first four-months of follow-up in the PIRP project; the left side of the graph shows the trend of admissions in the four years preceding PIRP enrolment; the right side shows the trend of admissions in the first four years of PIRP follow-up. The graphs include admissions for any diagnosis of patients with different status at 4 years since their enrolment in the PIRP project.

In stayers, a decrease of 90.15 admissions per 1,000 patients was observed in the first 4 months after enrolment in the PIRP compared to the previous quarter. In the 4 years preceding the first PIRP visit, their hospitalization trend increased by 10.73 AP4M, while the linear trend of AP4M in the first 4 years following enrolment in the PIRP increased by 2.64 units, thus indicating a net reduction of 8.09 AP4M. Sensitivity analysis conducted for stayers by splitting the period of enrolment into 3-year periods provided extremely consistent results (Figure 3).

**Figure 3:**
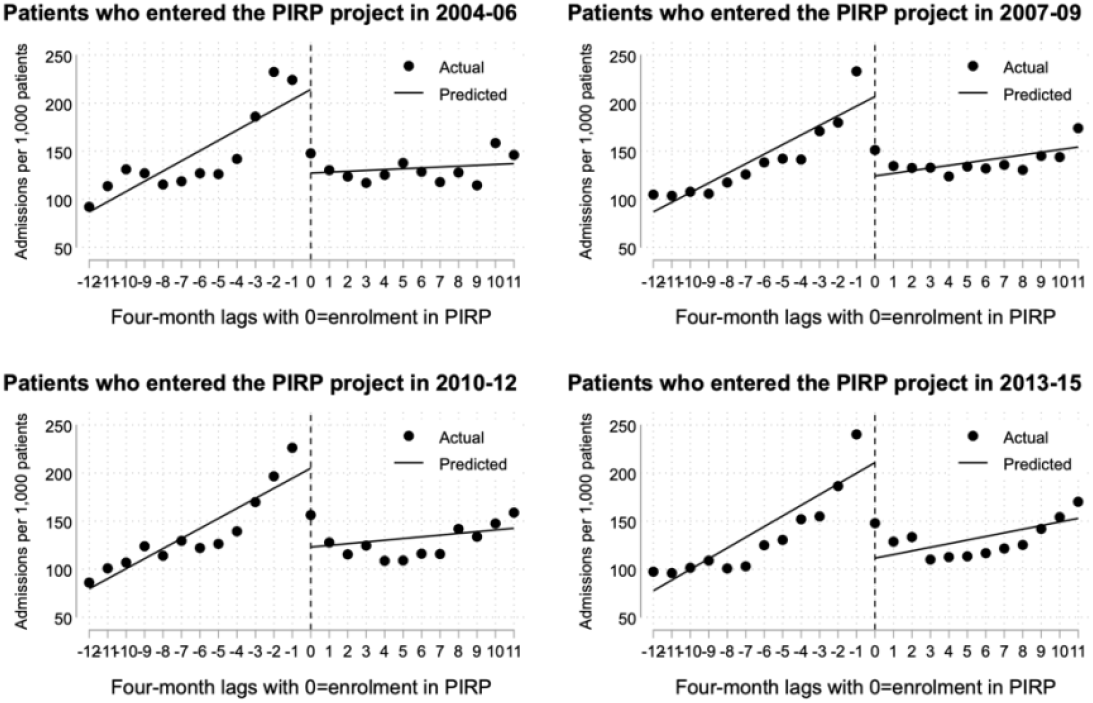
ITS of the number of hospitalizations by four-month periods, on different periods of enrolment in the PIRP project - only patients who maintained CKD-ND status at 4 years (stayers)

Subgroup analyses on survivors further showed that all main diagnoses had a significantly increasing trend in the number of hospitalizations in the 4 years before enrolment in PIRP (Figure 4, Table 4).

**Figure 4:**
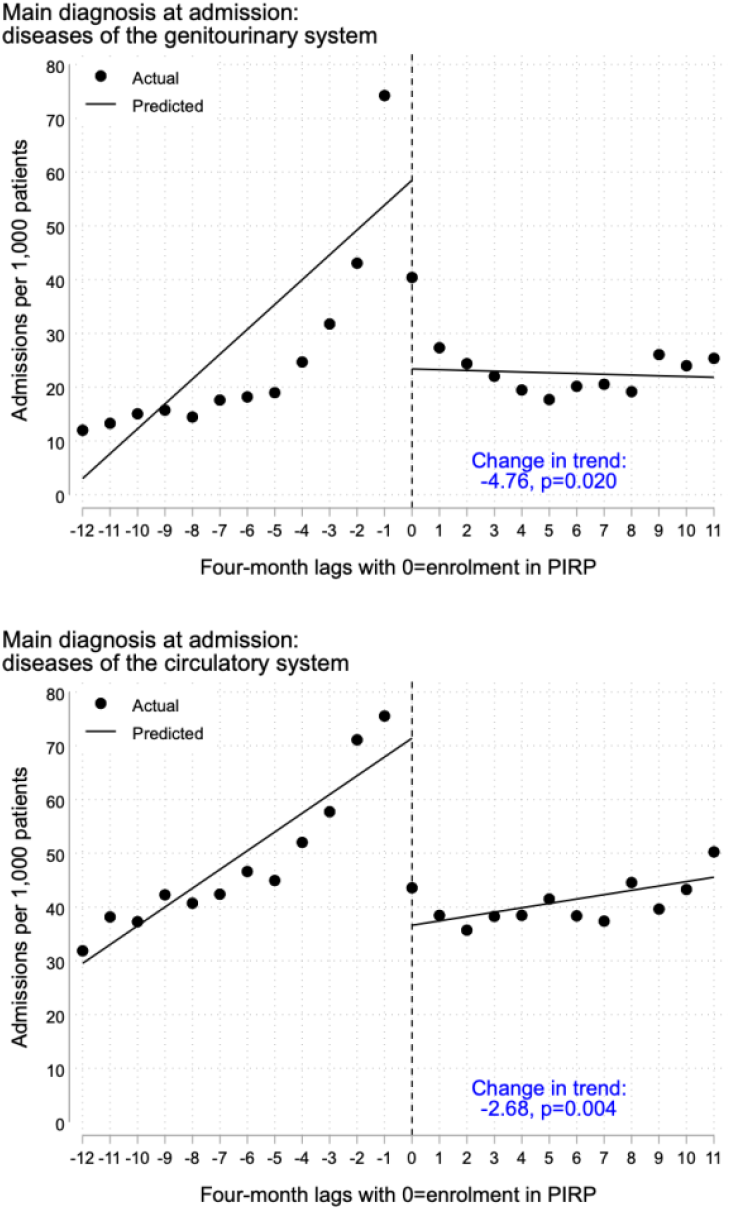
ITS of hospitalizations with diagnosis at admission of diseases of the genitourinary and circulatory systems, per 1,000 patients and four-month periods (stayer CKD-ND patients)

With the exception of diseases of the respiratory system and injuries, there was a large, or relatively large, generalized reduction in the number of hospitalizations soon after initiating PIRP follow-up.

Hospitalizations whose main diagnoses at discharge were disease of the circulatory system and disease of the genitourinary system showed the strongest increasing trend in the 4 years before PIRP enrolment, the largest gap and the sharpest decrease in the linear trend of hospitalizations after enrolment in PIRP (Figure 4). The enrolment in PIRP did not change the trend of hospitalizations for the other main diagnoses, except for the miscellaneous group of ‘other’ diagnoses, which significantly decreased (Table 4). A similar pattern was seen for patients with HF and CAD at PIRP enrolment (Figure 5 and Table 4), whose trend of hospitalizations was markedly increasing before being treated in PIRP and slowly increasing afterwards (or slowly decreasing in the case of admissions with CAD as main diagnosis). A significant increasing trend of hospitalizations prior PIRP enrolment and a significant drop at the first four-month period after enrolment was observed for every CKD stage (Table 4). The rate of hospitalization was higher with severity of the disease, and the change estimated in the hospitalizations’ trend was particularly marked for stages 5 (−37.06 from +41.27) and 4 (−10.88 from +14.21). As a result, a slight, non-significant (except for stage 3B) increase of AP4M was observed in the first four years of PIRP follow-up. In particular, patients who entered the PIRP project already in stage 5, and who could be followed up for at least 4 years, switched from a steeply increasing trend to one similar to those in stages 3B and 4, ranging from 2.77 to 4.21 AP4M.

**Figure 5:**
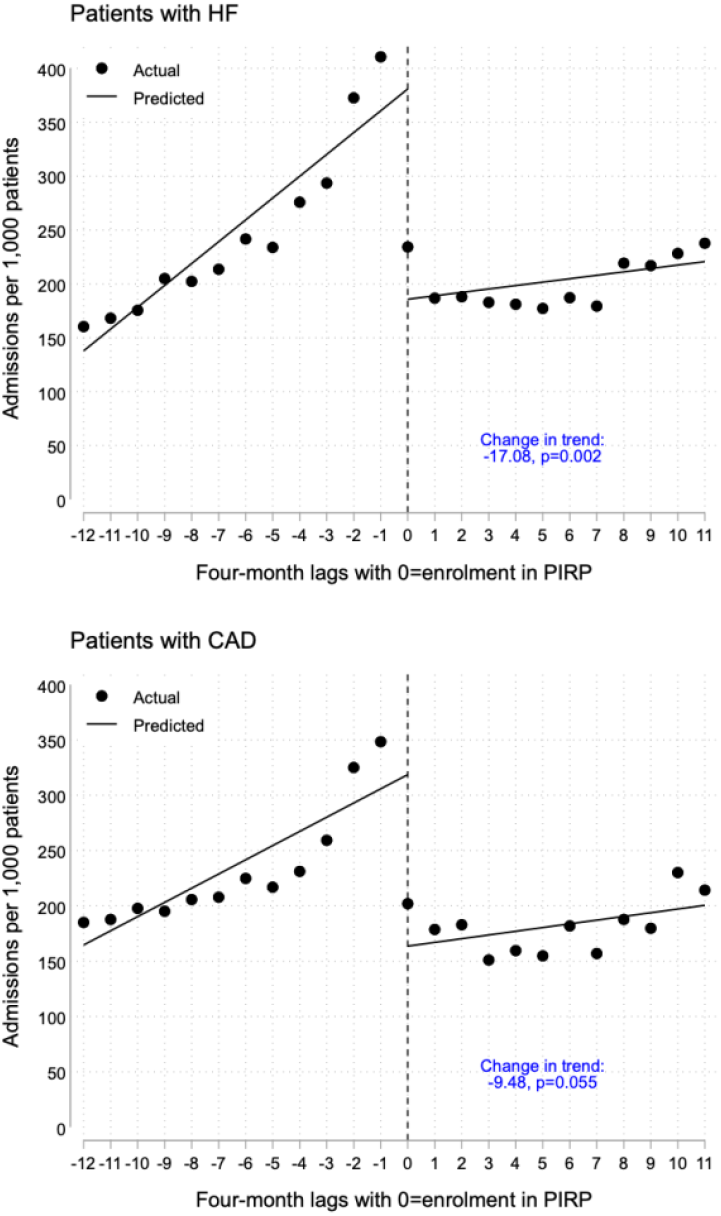
ITS of hospitalizations per 1,000 patients and four-month periods of stayer CKD-ND patients with heart failure (HF) and coronary artery disease (CAD)

With regard to treatment, 76.7% of patients were already under treatment with angiotensin-converting enzyme inhibitors and angiotensin receptor blockers (ACEi/ARBs), more than 50% were under treatment with beta-blockers, calcium-antagonists, diuretics, statins and low dose aspirin or clopidogrel and 11-12% were treated with aldosterone blockers and warfarin (Table 5). In the first year of follow-up in the PIRP project the proportion of patients under treatment increased in every main pharmacological group with the exception of ACEi/ARBs, that slightly reduced to 76.4%. Specifically, treatment with diuretics, ACEi/ARBs and calcium-antagonists was introduced in more than 10% of patients who were not previously treated with these drugs. Also, in proportions ranging from 1.4% to 8.3% (aldosterone blockers), treatment was interrupted soon after entering PIRP, in order to control hyperkalemia.

**Table 5:** Patients under treatment before entering the PIRP project and patients whose treatment was modified in the first year of follow-up. Main ATC groups for CKD.

| ATC group | Already under treatment* | Started treatment** | Interrupted treatment*** | Modified treatment* | In treatment in the first PIRP year* |
| --- | --- | --- | --- | --- | --- |
| ACEi/ARBs | 7804(76.7%) | 259(11.0%) | 298(3.8%) | 557(5.5%) | 8043(76.4%) |
| Aldosterone antagonists | 1087(10.7%) | 146(1.6%) | 90(8.3%) | 236(2.3%) | 1143(11.2%) |
| Beta blockers | 5093(50.1%) | 407(8.0%) | 84(1.6%) | 491(4.8%) | 5416(53.3%) |
| Calcium-antagonists | 5101(50.2%) | 517(10.2%) | 209(4.1%) | 726(7.1%) | 5409(53.2%) |
| Diuretics | 5184(51.0%) | 743(14.9%) | 73(1.4%) | 816(8.0%) | 5930(57.6%) |
| Statins | 5569(54.8%) | 442(9.6%) | 88(1.6%) | 530(5.2%) | 5923(58.2%) |
| Warfarin | 1245(12.2%) | 154(1.7%) | 44(3.5%) | 198(1.9%) | 1355(13.3%) |
| Low dose aspirin and clopidogrel | 5727(56.3%) | 357(8.0%) | 135(2.4%) | 492(4.8%) | 5949(58.5%) |

\* percentages calculated on all stayer patients (n=10169)
\*\* percentages calculated on stayer patients who were not under treatment before enrolment in PIRP
\*\*\* percentages calculated on stayer patients who were under treatment before enrolment in PIRP

## DISCUSSION

In this study, we evaluated the effect of the PIRP project (Prevenzione dell’Insufficienza Renale Progressiva) on hospital admissions in patients with progressive CKD. Patients who entered the PIRP project experienced a reduction in hospital admissions, in particular and in substantial manner hospitalizations due to cardiovascular diseases and genitourinary pathologies.

However, the hospitalization trend was not reduced in CKD-ND patients that in the 4 years following enrollment in the PIRP project did not survive or were forced to start dialysis. Most of these patients were already in an advanced stage of the disease at the moment of their enrolment in the PIRP project, and had a higher rate of comorbidities, previous major cardiovascular events and a significantly higher Charlson index, compared to “stayers” (patients who completed the same 4-year observation period). This finding reinforces the need to engage CKD-ND patients in a management program at an early stage of the disease, to maximize the benefits of appropriate surveillance: avoiding the progression of CKD and cardiovascular involvement. Consistently, the hospitalization rate remained higher with increasing stage of the disease.

In “stayers”, the observed abrupt and persistent reduction in the trend of genitourinary hospitalizations after enrollment in PIRP, could be interpreted as the result of better control of fluid overload, hyperkalemia and blood pressure, of the reduction of anti-inflammatory drugs and treatment of uncomplicated infections in an outpatient setting. The reduction of hospitalizations due to cardiovascular causes might be also attributed to the implementation of appropriate combination of pharmacotherapy (e.g. renin–angiotensin–aldosterone system blockers, beta-blockers, diuretics and statins), life styles modifications and a tailored therapy for heart failure or other cardiovascular complications.

Hospitalizations for cancer were also reduced, although the temporal trend did not differ after the entry in the PIRP project. It is unlikely that a nephrological intervention could affect the progression of neoplasms, therefore we hypothesize that patients who stayed 4 years in CKD-ND condition after entering the project were likely to have their cancer cured, and less needing to be hospitalized with this main diagnosis. Moreover, it cannot be ruled out that a change in the regional policy regarding tumor-related hospitalizations towards a wider use of day hospitals occurred during the study period. For respiratory problems and for infections, the reasons for the increase in terms of number of hospitalizations could be explained by the natural propensity of CKD patients to develop respiratory [18–19] and infectious complications [6], both difficult to prevent.

The result that we obtained in the CKD-ND stayer patients is even more relevant because it cannot be attributed to the utilization of drugs such as SGLT2 or MRAs antagonists, which proved to be extremely effective in the treatment of renal and cardiovascular diseases and consequently in reducing the related hospitalizations [20–22]. These drugs were not yet available in Italy during the time period considered in this study.

Several studies have shown that if the outpatient model of care is effective, the number of hospitalizations can be reduced [23]. Chronic kidney disease is included in the list of “Ambulatory Care-Sensitive Conditions (ACSC)”, those clinical conditions in which a large part of hospitalizations can be avoided through timely and effective outpatient care [7, 23–24]. This includes monitoring the disease, taking preventive measures, counteracting therapeutic inertia and providing lifestyle advice and patient education. In a recent paper, Chong et al. [7] demonstrated that a poor continuity of care in CKD patients was associated with a greater rate of a CKD-related ACSC.

A recently published pragmatic trial by Vasquez et al. [25] seems to counteract these hypotheses and our results, finding that patients randomized to receive usual care or an intervention including practice facilitators to assist primary care providers in delivering guideline-based interventions, showed similar hospitalization rates at 1 year. The intervention adopted in this study entails an exclusive GPs management of CKD patients, while in the PIRP project a relevant role is played by Nephrology specialists. The interventions provided in the PIRP project are not limited to implement the best therapies according to the Guidelines or strengthen patients’ adherence to therapy, but Nephrology specialists have treated on an outpatient setting some complications that would otherwise have led to hospital admissions. The guidelines play a crucial role in standardizing patient care but are not very suitable for a tailored application on single particular clinical cases.

Our study underlines that an approach based on nephrological clinical competence with some aspects shared with GPs is fundamental in reducing hospitalizations in CKD patients, both immediately after enrollment in the project and in the following 4 years, especially if they are intercepted in non-advanced stages. In fact, throughout the PIRP project, the outpatient infrastructure has been strengthened in all Nephrology Units of the Emilia Romagna Region, resulting in increased availability of outpatient access. The facilitated pathway and access favored a higher adherence of patients to the prescribed therapies, and allowed an earlier treatment in an outpatient setting for those complications that prior to the start of PIRP required a hospital admission. It should also be noted that the PIRP Project provides that, after each hospitalization for any cause, patients are reviewed in the nephrological outpatient clinic within a short period. In this way it was perhaps possible to prevent some re-hospitalizations. In fact, Doshi and Wish have demonstrated that an early nephrology consultation after discharge of patients with CKD may change the trends in 30-day readmissions and reduce the cost of care [26]. Furthermore, an early nephrological visit after discharge also allows to re-start effective therapies, which sometimes had to be suspended in the acute phase of hospitalization. However, the effectiveness of nephrologist’s interventions on hospital admissions does not emerge clearly from our work and only properly designed intervention studies could clarify a specific role.

Our study has several strengths, namely the large number of CKD-ND patients followed over several years; data on hospitalizations retrieved from official and reliable sources; the application of fairly homogeneous therapeutic management and follow-up protocols. Limitations include not estimating the balance between savings due to the reduction of hospitalizations and the overall costs of a CKD management program. Furthermore, the study did not include other patient-related factors that could influence the use and frequency of hospitalization, such as their socio-economic condition, site of residence and health literacy. In fact, a recent study in the CRIC cohort showed that adults with limited health literacy had a higher risk of CKD progression, cardiovascular events and hospitalizations [27]. Lastly, it is possible that our findings are not generalizable to other regions with different approaches in primary care delivery, or capacity to care for patients within the community.

### Conclusion

Our study shows that, by implementing an integrated public health project involving nephrologists and GPs, an optimal management and continuity of care is ensured to patients with CKD, also making possible to reduce preventable hospitalizations, and potentially leading to significant improvement in patients’ quality of life and healthcare cost savings.

## Data Availability

All data produced in the present study are available upon reasonable request to the authors

## ACKNOWLEDGEMENTS

We would like to thank all the patients and health professionals participating in the PIRP project.

## AUTHORS’ CONTRIBUTIONS

Research idea and study design: ASa, MM, DG; data acquisition: VA, AB, SC, GD, EF, MG, GLM, EM, RR, RS, ASt, AZ; data analysis/interpretation: ASa, MM, DG; statistical analysis: DG; supervision or mentorship: ASa, MM. Each author contributed important intellectual content during manuscript drafting or revision and agrees to be personally accountable for the individual’s own contribution and to ensure that questions pertaining to the accuracy or integrity of any portion of the work, even one in which the author was not directly involved, are appropriately investigated and resolved, including with documentation in the literature, if appropriate.

